# Deep Conformal Supervision: a comparative study

**DOI:** 10.1101/2024.03.28.24305008

**Authors:** Amir M. Vahdani, Shahriar Faghani

## Abstract

**Background:** Trustability is crucial for Al models in clinical settings. Conformal prediction as a robust uncertainty quantification framework has been receiving increasing attention as a valuable tool in improving model trustability. An area of active research is the method of non-conformity score calculation for conformal prediction.

**Method:** We propose deep conformal supervision (DCS) which leverages the intermediate outputs of deep supervision for non-conformity score calculation, via weighted averaging based on the inverse of mean calibration error for each stage. We benchmarked our method on two publicly available datasets focused on medical image classification; a pneumonia chest radiography dataset and a preprocessed version of the 2019 RSNA Intracranial Hemorrhage dataset.

**Results:** Our method achieved mean coverage errors of 16e-4 (CI: le-4, 41e-4) and 5e-4 (CI: le-4, 10e-4) compared to baseline mean coverage errors of 28e-4 (CI: 2e-4, 64e-4) and 21e-4 (CI: 8e-4, 3e-4) on the two datasets, respectively.

**Conclusion:** In this non-inferiority study, we observed that the baseline results of conformal prediction already exhibit small coverage errors. Our method shows a relative enhancement, particularly noticeable in scenarios involving smaller datasets or when considering smaller acceptable error levels, although this improvement is not statistically significant.

## Introduction

In contemporary medicine, the integration of artificial intelligence (Al) is rapidly expanding, offering innovative solutions for diagnosis, treatment, and patient care. Its adoption signifies a transformative shift in medical practice, driving advancements in precision medicine, clinical decision-making, and healthcare management.

However, model trustworthiness and explainability remain ongoing challenge, particularly in medical context, where decision-making is of tremendous implications. Uncertainty quantification (UQ) methods aim to provide a measure of a model’s uncertainty, representing the reliability of its predictions, or lack thereof. This is particularly crucial in clinical settings, where the accurate assessment of uncertainty ensures informed decision-making and patient safety.

Deep learning models conventionally consist of several ‘layers’ such as linear layers, activation layers, etc., performing a variety of mathematical operations arranged in sequential fashion (an initial input layer, hidden layers and a final output layer). Several approaches to UQ for these models are available, including (1):

- Probabilistic approaches: Utilize prior assumptions regarding the distribution of the data, and include ensemble methods, Bayesian methods, and evidential deep learning.
- Frequentist approaches: Focus on the data distribution within dataset, with no reliance on prior beliefs or assumptions regarding statistical distribution. Conformal prediction, explained below, is the most notable method in this category.

Conformal prediction (2) is a relatively novel approach in providing prediction sets/intervals with arbitrary guaranteed confidence levels of containing the correct target prediction. This framework has already been utilized in clinical research (3, 4), with noticeable potential due to its non-parametric modeling and robust generalizability to data previously unseen by the model. Of particular importance is the method of obtaining non-conformity scores in this approach, which are used to quantify the ‘strangeness’ of a particular subject from the model’s perspective, for which there are several options depending on the modeling scenario. Theoretically, this approach only needs exchangeability (i.e., invariance to the order of the data used to train the model and later data on which the model is expected to provide predictions), as compared to identical and independent distributions (i.i.e), which is the assumption required by many other uncertainty quantification approaches. In practice, a degree of deviation from the expected level of confidence will remain in previously unseen data, or in deep learning terms, test data. Asides from its robustness and alleviation of the need for distribution modeling (compared to approaches such as Bayesian uncertainty quantification), conformal prediction also has the attractive property of being a purely post hoc implementation, able to work with any statistical, machine learning or deep learning model without needing to change or retrain an already available model. We will provide a detailed explanation of conformal prediction in the setting of this research in the methods section.

Deep supervision (5), is a method that aims to obtain predictions from the hidden layers of the model alongside its output layer, aiming to use those intermediate outputs as additional inputs for the loss function (hence the name ‘deep’ supervision), providing benefits such as reduced training difficulty, robustness to noise and artifacts, and faster convergence (6). Additionally, the heterogeneity of the model’s several outputs can be used for UQ, similar to the ensemble methods mentioned above. This approach has already been used in clinical research utilizing deep learning models, such as breast cancer detection using breast ultrasound (7) and brain image semantic segmentation, e.g., brain tumors (8-10) and stroke lesions (11).

Starting with the intuition that utilizing the hidden layers of a deep learning model can lead to a better assessment of model uncertainty, in this work we propose a method of integrating conformal prediction in a deep supervision framework, titled deep conformal supervision (DCS). We benchmark our method on two public medical image classification datasets of different modalities, covering both binary and multiclass classification scenarios, and provide a comparison between this method and a conventional implementation of conformal prediction.

## Methods

### Datasets

We utilized two publicly available datasets for our experiments. The first dataset contains 5856 chest radiographs, from distinct subjects, 4273 with pneumonia and 1583 without pneumonia (12). The second dataset is a preprocessed version of the 2019 RSNA Intracranial Hemorrhage Detection Competition dataset (13, 14).

### Conformal Prediction

Formally, the aim of conformal prediction is to satisfy the following:

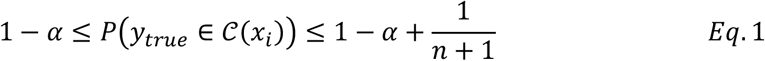

Where α represents an arbitrary error level (e.g., 0.05), y_true_ represents the ground truth label, *𝒞* (*x*_*i*_) is the prediction set, and *n* is the size of the calibration dataset. The calibration dataset is a distinct subset of the data available which should not be used during the training of the model, and is used to obtain non-conformity scores. Of note here is the distinction in the definition of calibration in the context of conformal prediction compared to its conventional definition in statistics.

Here is a short recipe for applying inductive conformal prediction (15) on a binary classification task:

1. Model training
  a. Dataset split into train, calibration and test subsets. Notably, cross-calibration is also an option, in a process akin to cross-validation.
  b. Train the model on the train subset
2. Calibration
  a. Calculation of non-conformity scores on the calibration subset. We used the hinge loss equation:

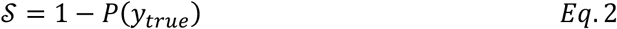

Where *P*(*y*_*true*_) represents the model’s predicted output for the ground truth label.
  b. Calculate the quantile corresponding to 1-α in the set of all the non-conformity scores obtained:

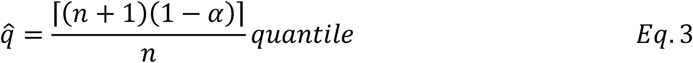

Where 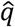 is the quantile threshold corresponding to the desired 1-α confidence level, taking into account finite sample correction.
3. Conformal prediction on the test set Calculate non-conformity scores per each possible class on the test dataset. Conformal prediction sets are constructed using the following condition:

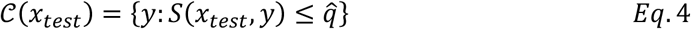

This would yield predictions in the space {[1], [0], [1, 0], ∅} (with 1 and 0 representing positive and negative labels, respectively), as compared to {0, 1}, yielded by conventional classification models using a threshold on the output probabilities.
4. Evaluation of conformal prediction on the test set. The two main metrics here are coverage (proportion of the sets containing the true label, which would ideally satisfy the condition in Eq. 1), and efficiency, which reflects how small the predicted sets are (in a range of [0-k], where k represents the number of classes).

### Model architecture

We used a pre-trained (on the ImageNet1K dataset (16)) ConvNext Tiny (17) backbone as our feature extractor. This backbone consists of 4 distinct stages, with a downscale operation between the stages. To implement deep supervision, we attached one multilayer perceptron (MLP) as a classifier ‘head’ to each stage (figure 1), yielding a tensor of shape [L, S] (where *L* is the number of labels, and *S* is the number of stages) as predictions for each subject. The detailed structure of the classifier head is available in the supplement material table 1. As we’ll see in the results sections, the earlier stages yield relatively inaccurate predictions, but we’ll leverage those to obtain better non-conformity scores, and thus, measures of uncertainty.

**Figure 1.**
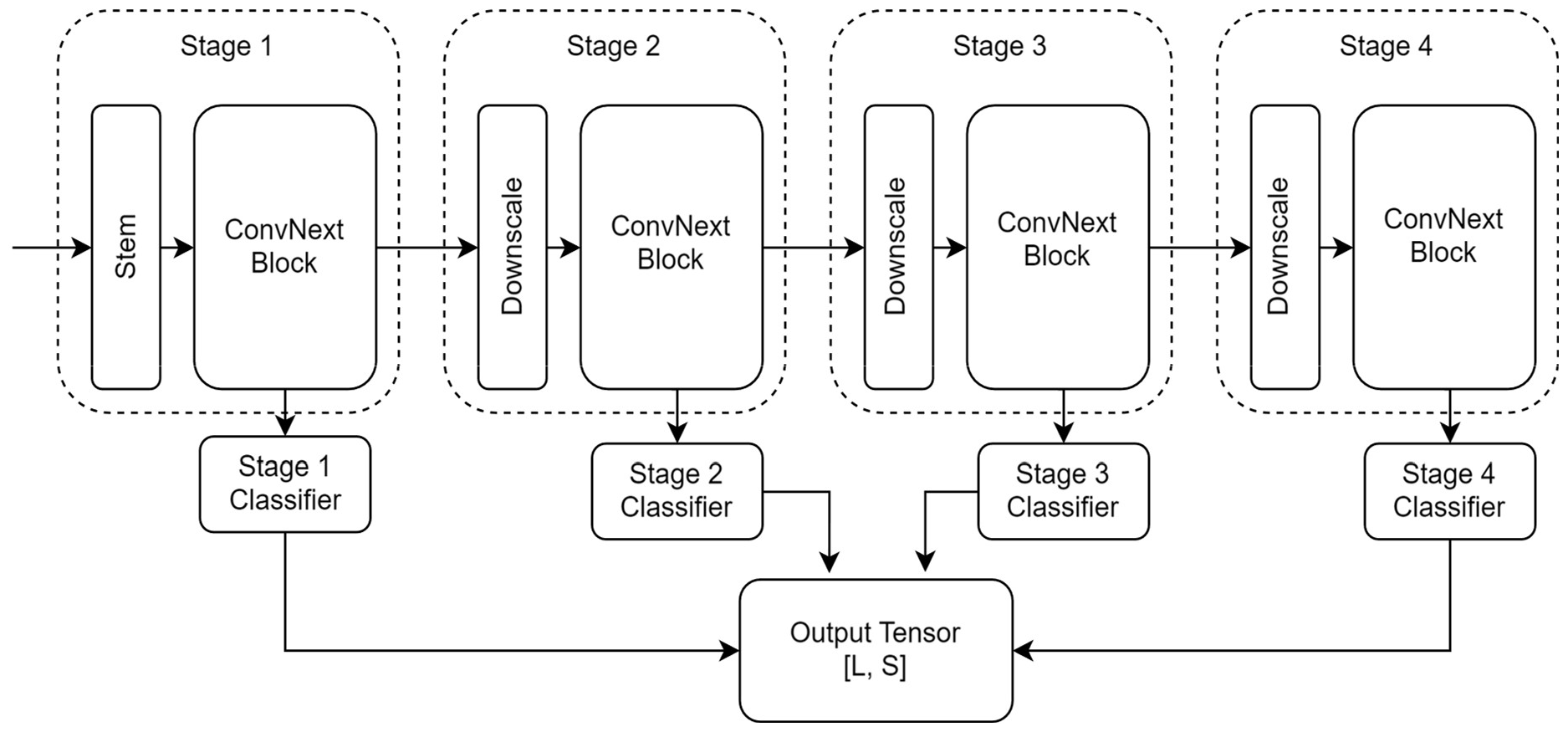
Overall model architecture

**Table 1.** Training Strategy.

| Table 1. Training Strategy |  |  |  |  |  |
| --- | --- | --- | --- | --- | --- |
|  | Pneumonia dataset (n=5856) |  | ICH dataset (n=970417) |  |  |
| Dataset split | Train + Calibration | Test | Train | Calibration | Test |
|  | 5-fold cross calibration | 674 | 82041 | n = 10000 | n = 5000 |
| Preprocessing | Resize to [768, 768] with padding, CLAHE, Normalization |  | Resize to [256, 256] with padding, CLAHE, Normalization |  |  |
| Data augmentation | RandRotate ($\pm 15^\circ$), RandShear ($\pm 2$), RandScale (0.95-1.05), RandHorizontalFlip, RandGaussianNoise ($\pm 5$) | | | | |
| Optimizer | AdamW (with AMSgrad, weight decay $\lambda=0.0001$) | | | | |
| Learning Rate | Cosine annealing LR scheduler with initial LR 0.001 |  |  |  |  |
| Batch Size | 16 (128 via gradient accumulation) |  | 256 (2048 via gradient accumulation) |  |  |
| Max Epochs | 100 (Backbone frozen during the 4 initial epochs) |  | 20 (Backbone frozen during the 4 initial epochs) |  |  |

**Table 2.** Pneumonia dataset classification metrics.

| Table 2. Pneumonia dataset classification metrics |  |  |  |  |
| --- | --- | --- | --- | --- |
| Model | Precision | Recall | F1 score | AUC |
| DCS stage 1 | 0.75 | 0.99 | 0.85 | 0.89 |
| DCS stage 2 | 0.96 | 0.93 | 0.94 | 0.95 |
| DCS stage 3 | 0.97 | 0.97 | 0.97 | 0.98 |
| DCS stage 4 | 0.97 | 0.97 | 0.97 | 0.98 |
| NDCS | 0.97 | 0.97 | 0.96 | 0.98 |

### Deep Conformal Supervision

Compared to a conventional implementation of conformal prediction in which *Eq*. 2 is used to obtain non-conformity scores, DCS outputs a vector of shape [1, S] (and non-conformity scores of the same shape) per label for each subject. Considering non-conformity scores have to be scalars in ℝ, an aggregation operation is needed. We tried several aggregation methods such as arithmetic mean, L2 norm etc. The best results were achieved via weighted averaging as follows:

1. Calculate mean calibration error per stage on the calibration subset:

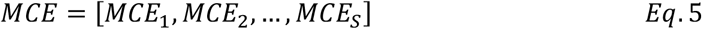
2. Calculate a weight vector according to the following:

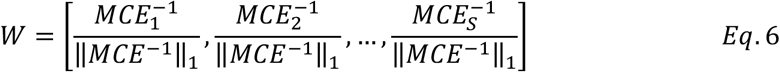

Where ∥*MCE*^-1^∥_1_ is the L1 norm of the reciprocal vector of MCE, used to scale W to unit magnitude.
3. Obtain non-conformity scores as follows:

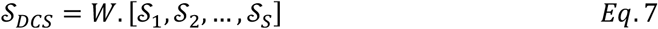

Where *𝒮* represents non-conformity scores obtained using Eq. 2. The rationale behind *Eq*. 7 is to better leverage the well-calibrated stages in obtaining non-conformity scores.

### Training Strategy

Random holdout test subsets were split first. Calibration was performed using 5-fold calibration on the Chest X-ray dataset (due to a relatively small dataset size) and single fold calibration on the MRI dataset. All splitting steps were stratified for class labels.

Preprocessing consisted of res121ng with padding, contrast limited adaptive histogram equalization (CLAHE) (18), and normalization using statistics obtained from the train subset.

Both models were trained using the AdamW optimizer, cosine annealing learning rate scheduler, and the binary cross-entropy loss function, with the backbone kept frozen for the initial 4 epochs. The calibration set was also used to validate the model during training to keep the model parameters with the best validation Fl score. Table 1 contains a detailed rundown of our training strategy for both datasets.

All the aforementioned steps (model design, training and evaluation) were implemented using PyTorch (19) v2.1.2. The Albumentations library (20) was also used for data augmentation.

## Results

### Classification results

Table 1 lists the validation metrics on the pneumonia dataset. We have listed metrics for DCS stages 1-4 along with the non-DCS (NDCS) model. As expected, classification metrics improve in subsequent deep supervision stages. The NDCS model achieves roughly similar metrics to DCS stage 4. Confusion matrices and calibration plots are available in the supplement material.

Table 3 lists the validation metrics on the ICH dataset (95% confidence interval across the 5 labels). Here, the trend of improvement across the stages is less consistent, especially across different labels. Confusion matrices and calibration plots are available in the supplement material.

**Table 3.** ICH dataset classification metrics.

| Table 3. ICH dataset classification metrics |  |  |  |  |
| --- | --- | --- | --- | --- |
| Model | Precision | Recall | F1 score | AUC |
| DCS stage 1 | [0.34, 0.88] | [0.25, 0.70] | [0.29, 0.77] | [0.77, 0.89] |
| DCS stage 2 | [0.37, 0.97] | [0.34, 0.90] | [0.36, 0.93] | [0.85, 0.96] |
| DCS stage 3 | [0.85, 0.89] | [0.69, 0.91] | [0.75, 0.90] | [0.94, 0.97] |
| DCS stage 4 | [0.84, 0.88] | [0.70, 0.92] | [0.76, 0.90] | [0.94, 0.97] |
| NDCS | [0.83, 0.89] | [0.69, 0.92] | [0.75, 0.90] | [0.94, 0.97] |

### DCS results

We valuated our conformal prediction procedure across a range of α values (0.001-0.1 for the CXR dataset and 0.001-0.15 for the MRI dataset) using three metrics:

- Coverage vs. ideal coverage: coverage on the test set compared to ideal coverage (1-α), across a range of α values (0.001-0.1 for the pneumonia dataset and 0.001-0.15 for the ICH dataset)
- Efficiency: average set size
- Coverage error (coverage - (1-α))

Two baselines were used for comparison with our proposed method (DCS):

- Calibration using only the outputs of the final stage of the deep supervision model (DCS stage 4)
- Calibration using the conventional model with no deep supervision (NDCS)

Figures 2 & 3 illustrate coverage and efficiency for the two datasets. Since all three methods achieved coverages very close to ideal coverage on the ICH dataset (attributable to its large sample size), we opted to visualize coverage error for this dataset instead of coverage.

**Figure 2.**
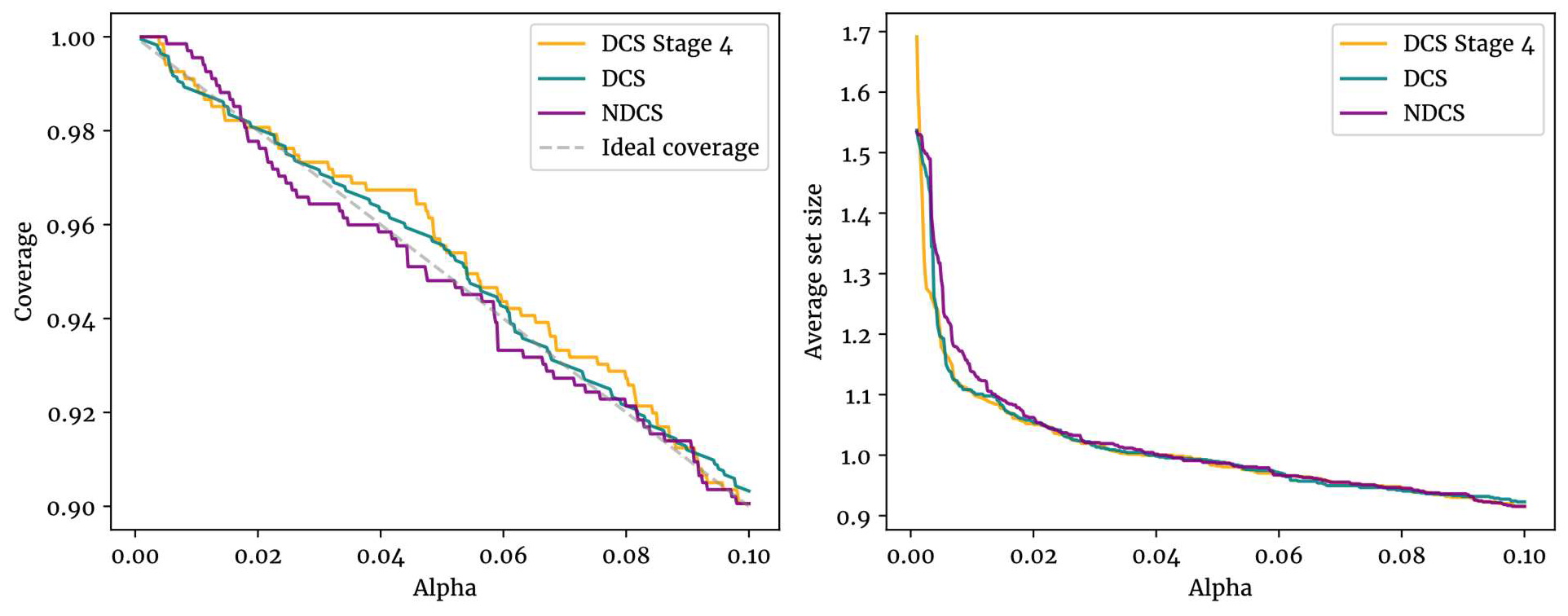
Conformal prediction metrics on the pneumonia dataset

**Figure 3.**
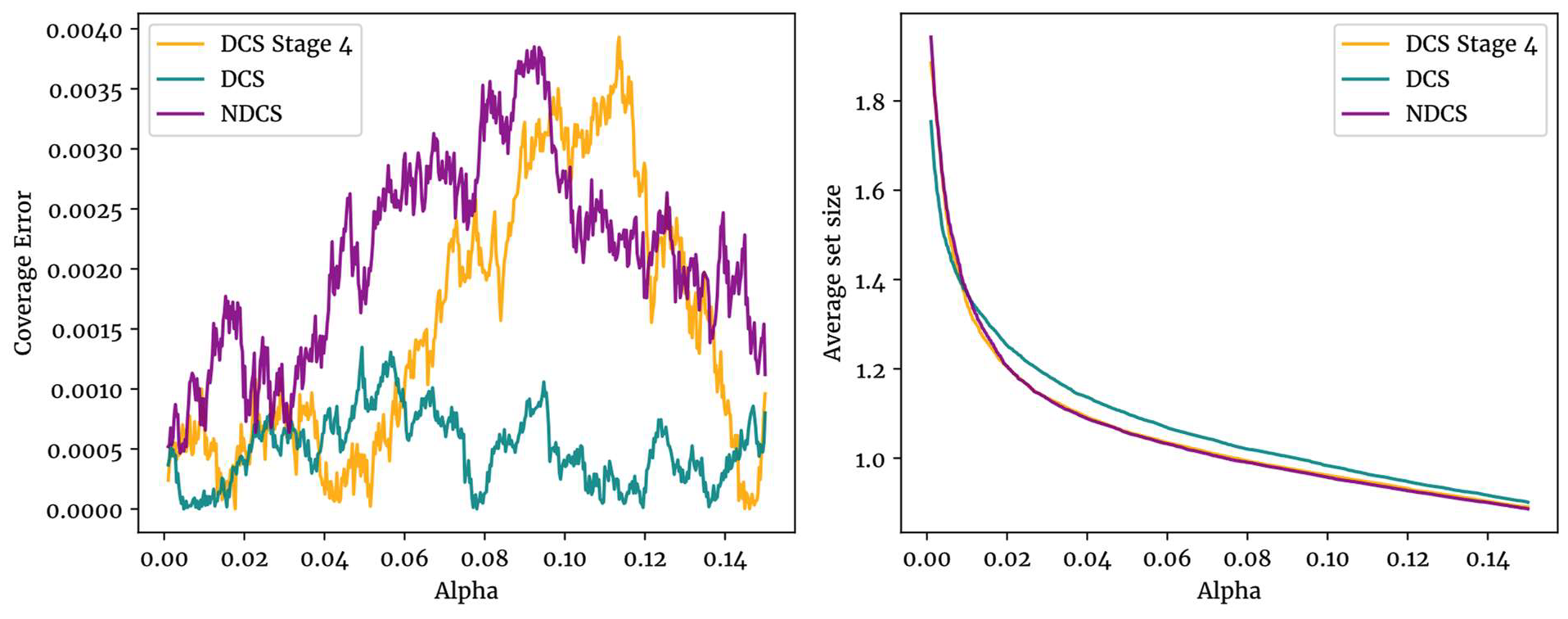
Conformal prediction metrics on the ICH dataset

Our proposed method achieves coverages closer to ideal on roughly the whole range alpha values used. It’s also worth noting that the DCS method has better efficiency when α is very small; the two baseline method basically fail on small values, predicting both classes (0 and 1) on almost all the test subjects.

As a final evaluation, we’ve also illustrated a box-and-whisker plot of coverage errors in figure 4. Our DCS method shows smaller mean coverage errors (16e-4 and 5e-4) compared to NDCS baselines of 28e-4 and 21e-4 on the two datasets, respectively. It also displays more stable test errors, with less changes in test error across the range of α values.

**Figure 4.**
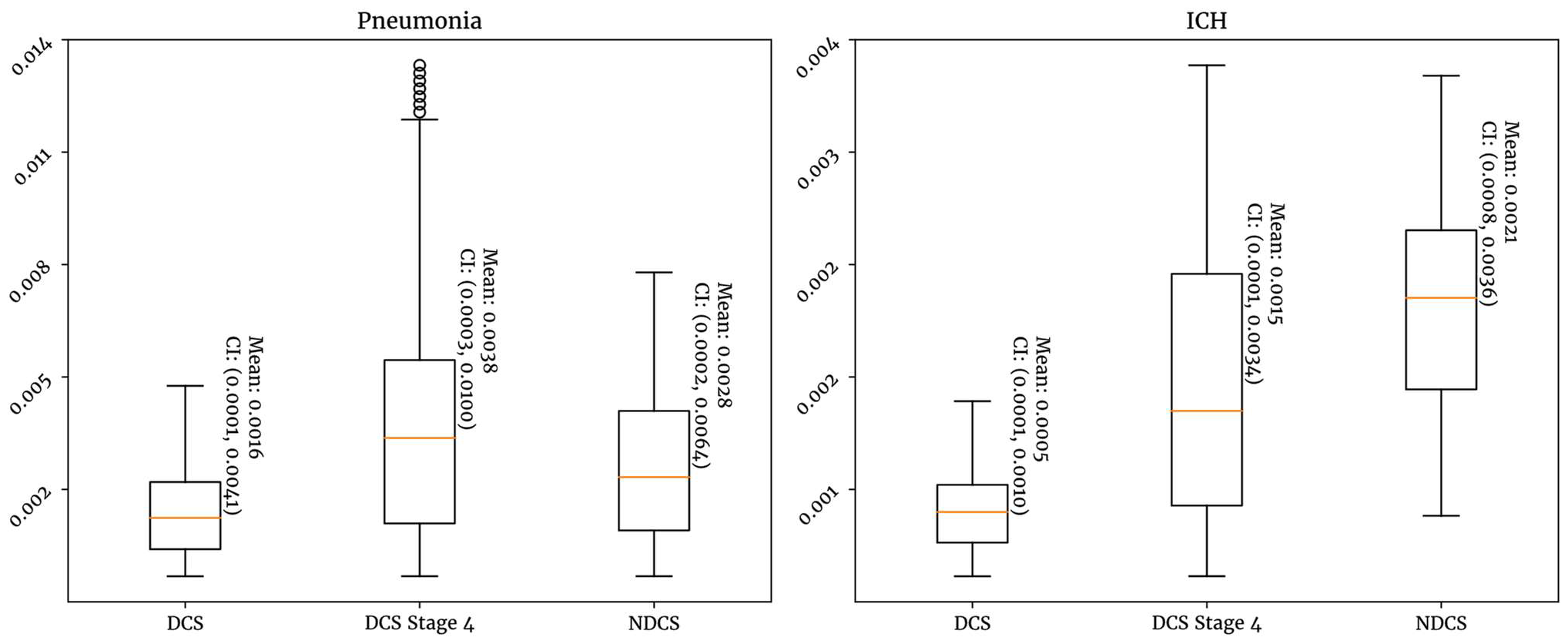
Coverage errors on both datasets. Mean coverage error and 95% confidence intervals are annotated.

## Discussion

Our findings indicate the potential for more robust implementations of conformal prediction when non-conformity score calculation methods better suited to deep learning model (such as our DCS method) are used. It’s worth noting that the baseline implementation of conformal prediction with no deep supervision achieves satisfactory metrics on the test set, but our proposed method still has merit particularly when high-stakes decision making is involved and errors are costly, which is the case in a lot of clinical scenarios; this improvement is more pronounced when dataset sizes are small (as in the case of the pneumonia dataset), which is of importance given the difficulty of assembling large datasets in clinical research.

The major limitation of our method is the compromise in the compatibility of conformal prediction with any (previously trained) model, since deep supervision requires modifications to both model architecture and training process.

To the best of our findings, tailoring the calculation of non-conformity scores to deep learning model is a relatively under-explored field (21). Future research could focus on improving the details of integrating deep supervision with conformal prediction, leverage the outputs of the hidden layers in a deep learning model, and/or integrate outlier detection alongside conformal prediction, as in (22).

## Conclusion

To summarize, our study demonstrates a novel method in non-conformity score calculation, with relatively improved coverage compared to conventional implementations, particularly when the dataset or alpha value for conformal prediction is small, with no decline in efficiency.

## Data Availability

All data produced are available online at:
https://kaggle.com/competitions/rsna-intracranial-hemorrhage-detection
https://www.kaggle.com/datasets/jhoward/rsna-hemorrhage-jpg

https://kaggle.com/competitions/rsna-intracranial-hemorrhage-detection

https://www.kaggle.com/datasets/jhoward/rsna-hemorrhage-jpg

## Conflict of Interest

The authors have no conflicts of interest to disclose.

## Author Contributions

Conceptualization: all authors. Supervision: Shahriar Faghani. Writing-original draft: Amir M. Vahdani. Writing-review & editing: all authors.

---

**Supplementary figure 1.**
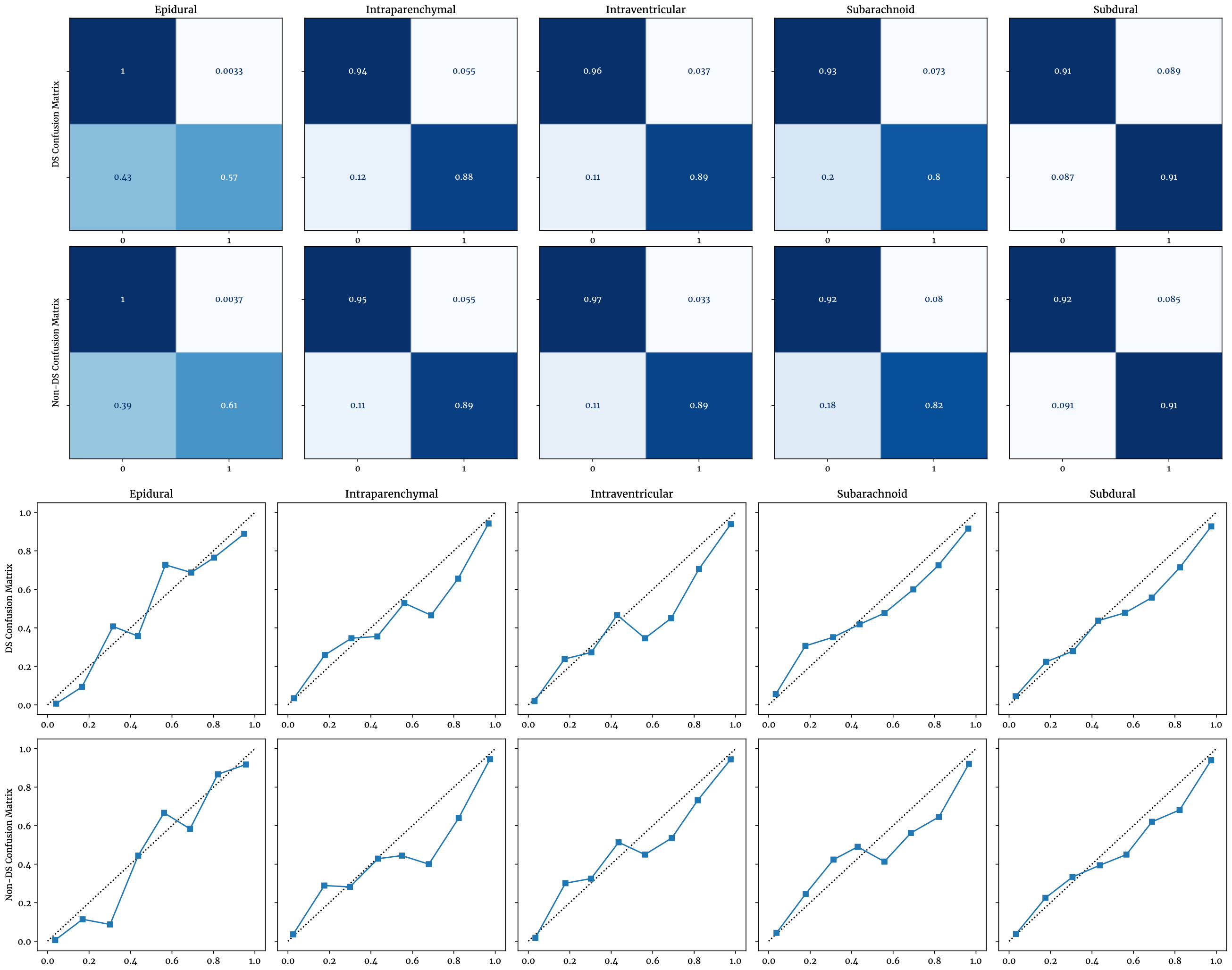
ICH dataset confusion matrices and calibration plots

**Supplementary figure 2.**
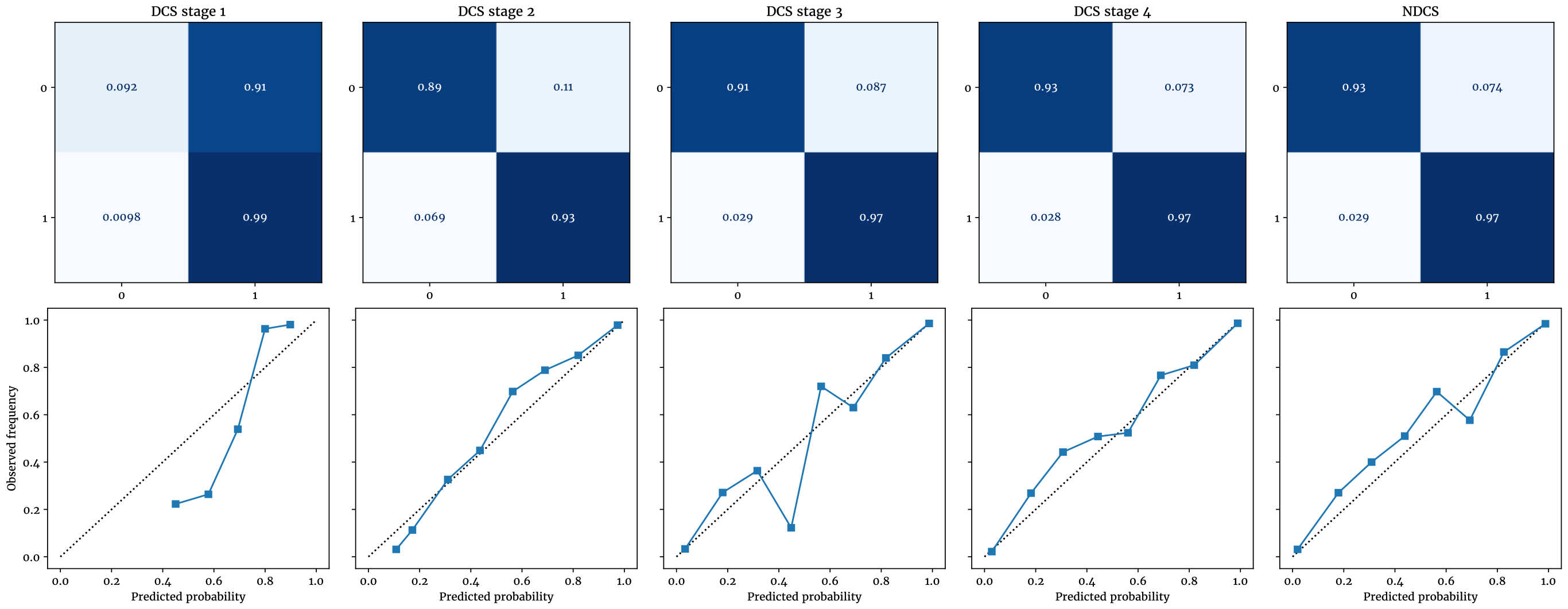
Pneumonia dataset confusion matrices and calibration plots

## Notes

### Competing Interest Statement

The authors have declared no competing interest.

### Funding Statement

This study did not receive any funding.

